# Understanding Bladder Cancer Screening Limits Through Comparative Modeling: The Maximum Clinical Incidence Reduction (MCLIR) Methodology

**DOI:** 10.64898/2026.01.27.26344939

**Authors:** Hawre Jalal, Stella K. Kang, Fernando Alarid-Escudero, Stavroula A. Chrysanthopoulou, David Ulises Garibay-Trevino, Bruce L. Jacobs, Karen M. Kuntz, Praveen Kumar, Jonah H. Popp, Yuliia Sereda, Mutita Siriruchatanon, John B. Wong, Thomas A. Trikalinos, CISNET Bladder Cancer Modeling Investigators

## Abstract

**Background:** Bladder cancer imposes substantial clinical and economic burden, yet key natural-history quantities that determine the potential effectiveness of screening—such as the size of the screen-detectable preclinical reservoir and the preclinical sojourn time—are largely unobservable. The Cancer Intervention and Surveillance Modeling Network (CISNET) uses standardized stress tests to compare independently developed microsimulation models and to clarify how differences in model structure translate into differences in intervention impact. The Maximum Clinical Incidence Reduction (MCLIR) framework estimates the maximum achievable reduction in clinically detected incidence following a one-time, perfect screening intervention, while Realistic Clinical Incidence Reduction (RCLIR) relaxes the perfect-test and/or perfect-treatment assumptions.

**Methods:** We applied the CISNET MCLIR/RCLIR protocol to three independently developed bladder cancer microsimulation models (COBRAS, Kystis, and SCOUT), each calibrated to common U.S. epidemiologic targets. We simulated a U.S. birth cohort born in 1950 and compared: (1) no-screening baseline; (2) one-time perfect screening with universally curative treatment (MCLIR) at ages 60, 65, 70, and 75; and (3) one-time realistic screening with cystoscopy sensitivity of 80% and perfect treatment (RCLIR-1) or usual-care treatment effectiveness (RCLIR-2). Outcomes included age-specific clinical incidence and incidence-reduction curves relative to baseline, as well as cumulative reductions over follow-up.

**Results:** Across models, median ages at first lesion emergence, clinical diagnosis, and onset of muscle-invasive disease were similar, but preclinical sojourn time differed meaningfully: COBRAS produced the shortest median sojourn time (2.1 years) compared with Kystis (3.3 years) and SCOUT (3.1 years). Under MCLIR, all models predicted an immediate drop in clinical incidence followed by attenuation toward zero as new lesions emerged after the screening age. Peak MCLIR at age 65 among White men ranged from 20% (COBRAS) to 21% (Kystis) and 32% (SCOUT), with reductions dissipating within ∼8 years in COBRAS and persisting ∼10 years in Kystis and SCOUT. In COBRAS and Kystis, incidence reductions later became negative, consistent with rebound emergence of new lesions among individuals whose first lesions were removed by screening. Under RCLIR-1, peak reductions were smaller and varied substantially across models (13%, 4%, and 37% for COBRAS, Kystis, and SCOUT, respectively). RCLIR-2 further reduced gains and produced more gradual decay, reflecting incomplete prevention under usual-care treatment. Across models, most residual post-screening incidence was attributable to new lesion emergence rather than missed detection or incomplete treatment.

**Conclusions:** In a standardized CISNET stress test, three bladder cancer microsimulation models imply a relatively short detectable preclinical phase, placing a modest upper bound on the effectiveness of one-time screening in the general population. Differences in MCLIR/RCLIR magnitude and persistence are explained by differences in implied sojourn time and detectable reservoir size. These findings motivate evaluation of risk-targeted and repeated early-detection strategies and highlight key empirical priorities for improving inference on bladder cancer natural history.

## Introduction

Bladder cancer remains a major and growing public-health challenge. It is the 6th most commonly diagnosed cancer in the United States, with an estimated 83,190 new cases in 2024, and the 8th leading cause of cancer mortality in men.^1,2^ Nearly 75% of cases are detected as non– muscle invasive disease, which, despite its relatively favorable prognosis, requires lifelong surveillance and repeated treatment due to high recurrence risk, resulting in an annual economic burden of approximately 6.5 billion dollars in the US alone.^3,4^ Even though exposure to major established risk factors such as cigarette smoking and environmental carcinogens has declined over time,^5,6^ population trends in bladder cancer incidence and mortality remained largely stable through the mid-2000s, with only a modest 1–1.5% annual decline in age-standardized incidence observed since 2005.^7^ In combination with demographic aging, these patterns suggest that the absolute number of bladder cancer cases will remain substantial, sustaining its clinical and economic burden.^8^

A central obstacle to prevention and early-detection policy in bladder cancer is incomplete knowledge of its natural history. Key events such as lesion initiation, the onset of a screen-detectable preclinical phase, progression to muscle-invasive disease, and the sojourn time from detectability to clinical diagnosis are largely unobservable in routine data. Mathematical microsimulation models are therefore useful for synthesizing epidemiologic and clinical evidence, inferring unobserved natural-history quantities, and projecting outcomes under counterfactual interventions. However, because different models may rely on different structural assumptions while still reproducing observed incidence patterns, comparative modeling is necessary to understand which assumptions drive divergent policy predictions and to identify robust conclusions. This comparative-modeling approach is a hallmark of the Cancer Intervention and Surveillance Modeling Network (CISNET), where multiple independently developed models are calibrated to common targets and then interrogated using shared stress tests.

The Maximum Clinical Incidence Reduction (MCLIR) analysis is one such CISNET stress test designed to reveal how models’ implicit natural-history assumptions constrain the potential effectiveness of screening. Conceptually, MCLIR asks: if a one-time, perfect screening examination were performed at a specific age, detecting and completely removing all existing detectable preclinical disease with perfect treatment, how much would subsequent *clinically detected* cancer incidence fall in the absence of any further screening? ^9-11^. The resulting reduction represents the maximum achievable decline in clinical incidence that is attributable solely to the reservoir and sojourn time of detectable preclinical disease at that age. Models with longer preclinical sojourn times or larger detectable reservoirs yield larger and more sustained MCLIR curves, whereas models with short sojourn times yield smaller, rapidly diminishing reductions. In this way, MCLIR provides an output-based, model-agnostic summary of natural history that is directly interpretable in screening terms and has proven useful in lung, cervical, breast, and colorectal CISNET comparisons ^9-11^.

A complementary metric, the Realistic Clinical Incidence Reduction (RCLIR), relaxes the “perfect test” assumption by applying a one-time screening scenario with empirically plausible sensitivity (and, in some applications, realistic downstream effectiveness), while still withholding any subsequent screening. By contrasting MCLIR with RCLIR, investigators can separate limitations imposed by the disease’s natural history from those imposed by imperfect detection and management. Together, the paired MCLIR/RCLIR framework offers a transparent way to decompose why screening may fail to prevent cancers, because lesions arose after screening, were missed at screening, or were detected but not successfully averted, and to compare those mechanisms across models ^10,11^.

We apply MCLIR and RCLIR analyses to three independently developed CISNET bladder cancer microsimulation models that have been calibrated to common U.S. epidemiologic targets but differ in key structural representations of lesion initiation, growth, detection, progression, and risk-factor effects.^8^ The models are Cancer of the Bladder R-based Analytic Simulator (COBRAS), Kystis, and the Simulation of Cancers of the Urinary Tract (SCOUT). The goal is to leverage their differences to clarify what each implies about the timing and duration of detectable preclinical bladder cancer, and therefore about the theoretical ceiling on screening benefit. By situating bladder cancer within the established CISNET stress-testing paradigm, this study provides (i) a standardized, screening-relevant description of natural-history assumptions across models, (ii) insights into why projected screening effects may diverge, and (iii) an evidentiary foundation for subsequent, policy-focused evaluations of candidate early-detection strategies in high-risk populations.

## Methods

We conducted a CISNET comparative model stress test using the Maximum Clinical Incidence Reduction (MCLIR) and Realistic Clinical Incidence Reduction (RCLIR) framework to characterize and compare the natural-history assumptions embedded within three independently developed bladder cancer microsimulation models. The stress test follows the standardized CISNET “Compare” protocol for MCLIR/RCLIR analyses used previously in other cancer sites^10-12^.

All models were run under harmonized population inputs and prespecified screening/treatment counterfactuals. We simulated a single U.S. birth cohort born in 1950 and contrasted: (1) a baseline with no screening, (2) a one-time *perfect* screening and curative treatment scenario (MCLIR), and (3) two one-time *realistic* cystoscopy screening scenarios (RCLIR-1 and RCLIR-2). The primary outputs were age-specific clinical incidence trajectories and their reductions relative to the no-screening baseline.

The three models (COBRAS, Kystis, and SCOUT) are population microsimulation models developed within the CISNET Bladder Working Group. The details of these models are described elsewhere^8^. In summary, each simulates individual life histories from birth to death, including bladder cancer initiation, progression, symptom onset/clinical detection, treatment, recurrence, and cause-specific mortality. Models were independently developed and calibrated to common U.S. epidemiologic targets, but differ in structure and parameterization, allowing comparative inference about unobservable natural history.^8^

COBRAS is a continuous-time discrete-event simulation that generates U.S. birth cohorts and tracks individuals through tumor initiation, growth/progression across non–muscle invasive and muscle-invasive pathways, and detection either by symptoms or incidental workup. Risk of onset and progression is modified by demographic factors and smoking histories. The model is designed for evaluating prevention, early detection, surveillance, and treatment strategies at the population level.

Kystis is also a continuous-time discrete-event microsimulation model of bladder cancer natural history in the U.S. population. It represents disease initiation, preclinical evolution, symptom development, clinical diagnosis, treatment, recurrence, and survival, and can be run as stacked birth cohorts to reproduce national trends.

SCOUT is a state-transition microsimulation model operating in discrete monthly cycles. It simulates U.S. birth cohorts forward in time, capturing transitions among health states corresponding to preclinical and clinically detected bladder cancer stages/grades, treatment, recurrence, and death. Like the other models, SCOUT is calibrated to U.S. incidence and survival patterns but uses a different temporal resolution and state-based representation.

Across all models, calibrated natural-history modules are overlaid with intervention modules (screening, treatment, surveillance), consistent with CISNET modeling principles. Following the Compare MCLIR protocol, each model simulated a single U.S. birth cohort centered on individuals born in the 1950s (the 1950 birth cohort in implementation), stratified by sex and race/ethnicity using harmonized U.S. life tables and bladder cancer risk-factor histories, such as the Smoking History Generator.^13^ Cohorts were simulated from birth until death without migration.

### Stress-test scenarios

1. *No-screening baseline*. In the baseline scenario, individuals progressed according to each model’s calibrated natural history with detection occurring only through symptoms or incidental clinical presentation (i.e., no organized screening). This scenario provides the counterfactual clinical incidence trajectory against which incidence reductions were computed.
2. *MCLIR scenario (one-time perfect screening + perfect treatment)*. To estimate maximum clinical incidence reduction, we imposed a one-time screening at ages 60, 65, 70 and 75 among individuals not yet clinically detected. Screening was assumed error-free (100% sensitivity and 100% specificity) for detecting all screen-detectable preclinical bladder cancers present at each age. All screen-detected cancers received immediate and universally curative treatment, eliminating subsequent clinical presentation from those lesions. No further screening occurred the screening age.
3. *RCLIR-1 scenario (realistic cystoscopy sensitivity + perfect treatment)*. RCLIR-1 replaces the perfect test with a plausible, one-time screen (e.g., cystoscopy) at age 65. Test specificity was assumed to be 100%, and sensitivity was set to an evidence-based value (80% in the base case), consistent with the high sensitivity reported for conventional white-light cystoscopy in diagnostic settings ^14^. Screen-detected cancers received immediate universally curative treatment. As in MCLIR, no repeat screening was allowed.
4. *RCLIR-2 scenario (realistic cystoscopy sensitivity + usual-care treatment effectiveness)*. RCLIR-2 used the same one-time cystoscopy screen at each age and the same sensitivity/specificity assumptions as RCLIR-1. However, screen-detected cancers were treated under usual-care effectiveness: treatment began immediately after screen detection and conferred the stage-specific survival and recurrence outcomes already embedded in each model’s calibrated treatment module (i.e., no universal cure). We assumed that treatment would cure 90% of lesions immediately. This scenario isolates the combined effect of imperfect detection and imperfect treatment.

### Outcomes

For each model and scenario, we estimated:

1. Age-specific clinical incidence rates (overall bladder cancer), defined as cancers that would present clinically after symptom onset or incidental diagnosis.
2. Incidence reduction curves under MCLIR and RCLIR relative to baseline:

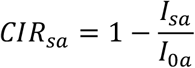

where *I*_*sa*_ is the clinical incidence at age a under scenario s and *I*_0*a*_ is baseline incidence at age a. These curves quantify the fraction of clinical incidence preventable by a single screen at a specified age under each set of assumptions ^10,11^.
3. Cumulative clinical incidence reduction over follow-up horizons (e.g., to age 100), computed as the integral of incidence reductions over age.

### Implementation and comparison across models

Each modeling group implemented the common scenarios using their native code, ensuring that the only differences across models arose from underlying natural history and treatment structure. Outputs were standardized to a common set of age bins and follow-up windows, then compared descriptively across models. Consistent with CISNET stress-test practice, we emphasize cross-model contrasts in MCLIR/RCLIR magnitudes, timing, and persistence as indicators of differences in (i) the size of the screen-detectable preclinical reservoir at each age and (ii) preclinical sojourn times leading to clinical diagnosis. ^11,12^.

## Results

The inferred timing of key natural-history events was comparable across models but with meaningful differences in preclinical sojourn time. For white men in the 1950 cohort, median age at first lesion emergence was 71–74 years, median age at clinical diagnosis 74–77 years, and median age at onset of MIBC 75–77 years (Table 1). The emergent preclinical sojourn time (the interval from becoming screen detectable to clinical diagnosis) was shortest in COBRAS (median 2.1 years) and longer in Kystis and SCOUT (medians 3.3 and 3.1 years, respectively), with wide interquartile ranges in all models (Table 1). These differences imply that a one-time screen would intercept a smaller detectable reservoir in COBRAS than in Kystis or SCOUT, and that any benefit should decay faster when preclinical sojourn times are shorter.

**Table 1.**
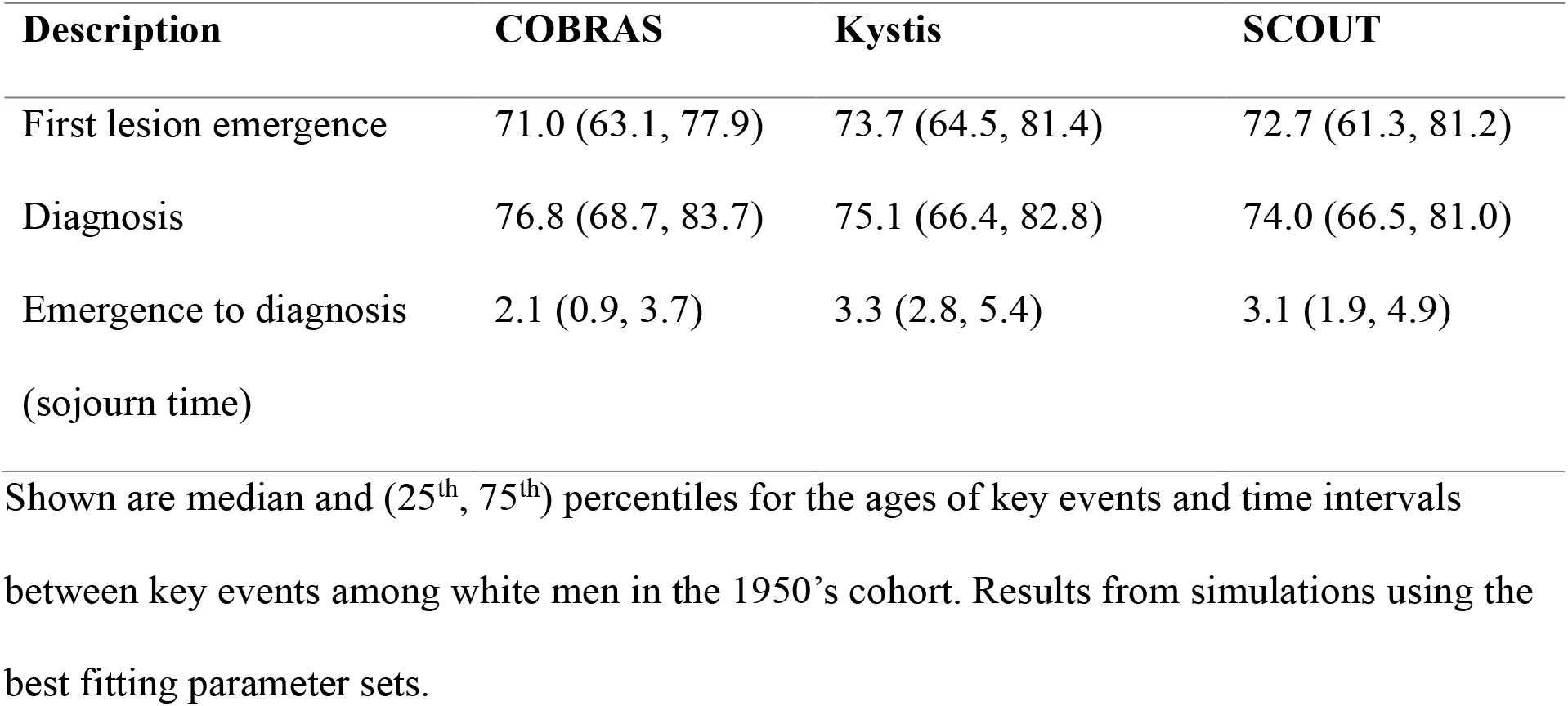
Ages of and time intervals between key events with each model (white men, 1950 cohort)

| **Description** | **COBRAS** | **Kystis** | **SCOUT** |
| --- | --- | --- | --- |
| First lesion emergence | 71.0 (63.1, 77.9) | 73.7 (64.5, 81.4) | 72.7 (61.3, 81.2) |
| Diagnosis | 76.8 (68.7, 83.7) | 75.1 (66.4, 82.8) | 74.0 (66.5, 81.0) |
| Emergence to diagnosis<br>(sojourn time) | 2.1 (0.9, 3.7) | 3.3 (2.8, 5.4) | 3.1 (1.9, 4.9) |

Shown are median and (25<sup>th</sup>, 75<sup>th</sup>) percentiles for the ages of key events and time intervals between key events among white men in the 1950's cohort. Results from simulations using the best fitting parameter sets.

### Maximum Clinical Incidence Reduction (MCLIR)

Figure 1 shows the age-specific clinical incidence reduction produced by a one-time perfect screen with universally curative treatment at ages 60, 65, 70 and 75, for men and women, and Blacks and Whites. All models predicted an immediate drop in clinical incidence, followed by gradual attenuation as new lesions emerged after the screening age.

**Figure 1.**
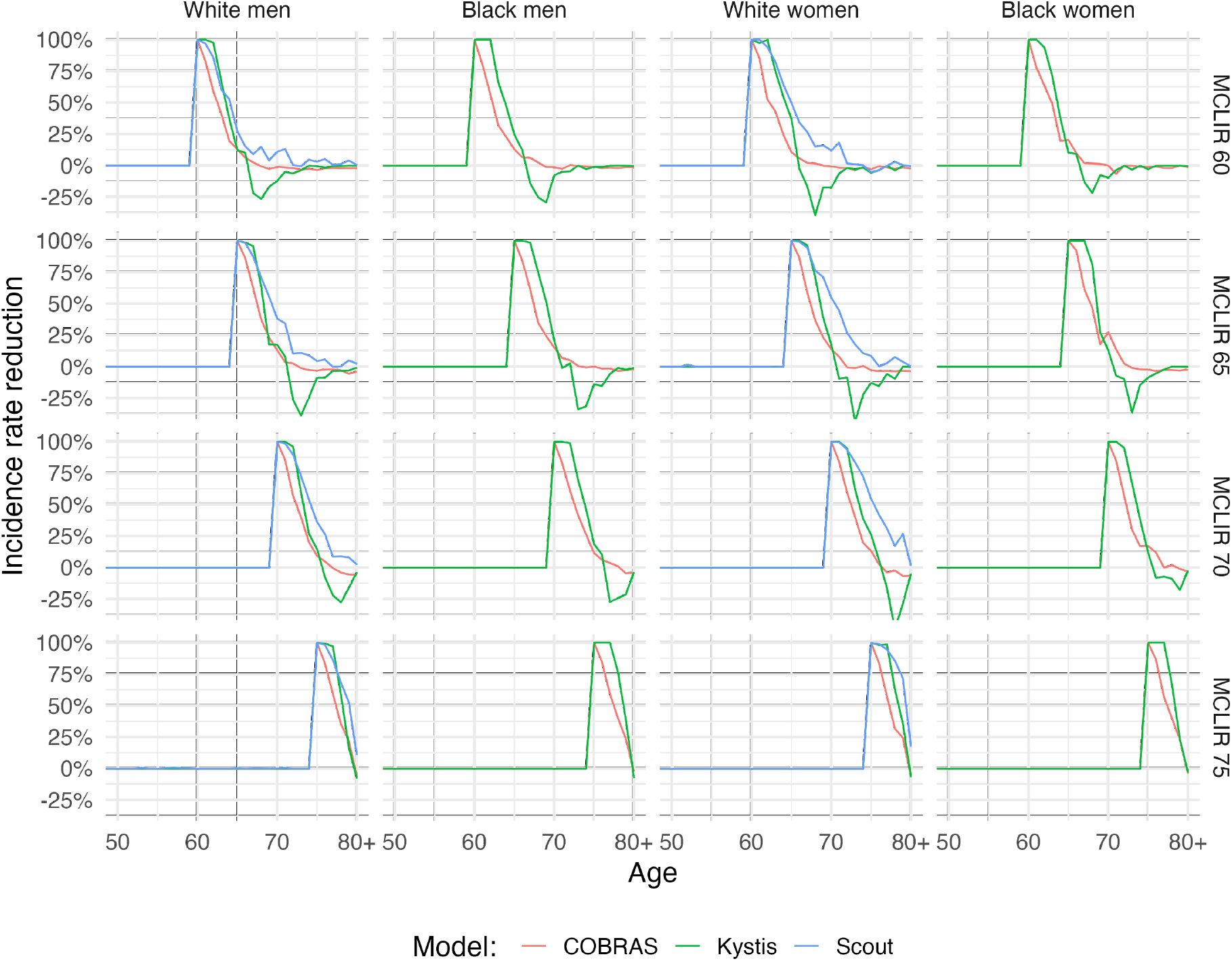
MCLIR curves after one-time perfect screening by age, sex and race

The maximum (peak) clinical incidence reduction occurred after screening in all models but differed in size (Figure 1). For example, COBRAS showed the smallest peak MCLIR65 among White men (peak reduction = 20% at age ∼70), whereas Kystis and SCOUT yielded larger peaks (Kystis: 21%; SCOUT: 32%). These results were consistent for other ages of MCLIR and sex and race categories. In addition, the MCLIR curves converged toward zero at older ages, but the speed of convergence differed (Figure 1). In COBRAS, reductions diminished rapidly and were near zero in about 8 years, consistent with its shorter sojourn time (Table 1). Kystis and SCOUT showed more persistent reductions of 10 years, consistent with their longer detectable preclinical sojourn times. Furthermore, in Kystis and COBRAS, the incidence reductions become negative. This is consistent with the emergent of new lesions among those in whom their first lesions were removed by the MCLIR intervention. These negative rebounds are more pronounced in the Kystis model.

### RCLIR-1 (realistic cystoscopy sensitivity, perfect cure)

Substituting realistic cystoscopy sensitivity reduced the immediate post-screening benefit in all models (Figure 2). For example, peak reductions for RCLIR-165 fell to 13%, 4%, and 37% in COBRAS, Kystis, and SCOUT, respectively. The proportional gap between MCLIR and RCLIR-1 was similar across models, indicating that differences in achievable benefit were driven primarily by natural-history structure rather than by model-specific implementation of test sensitivity.

**Figure 2.**
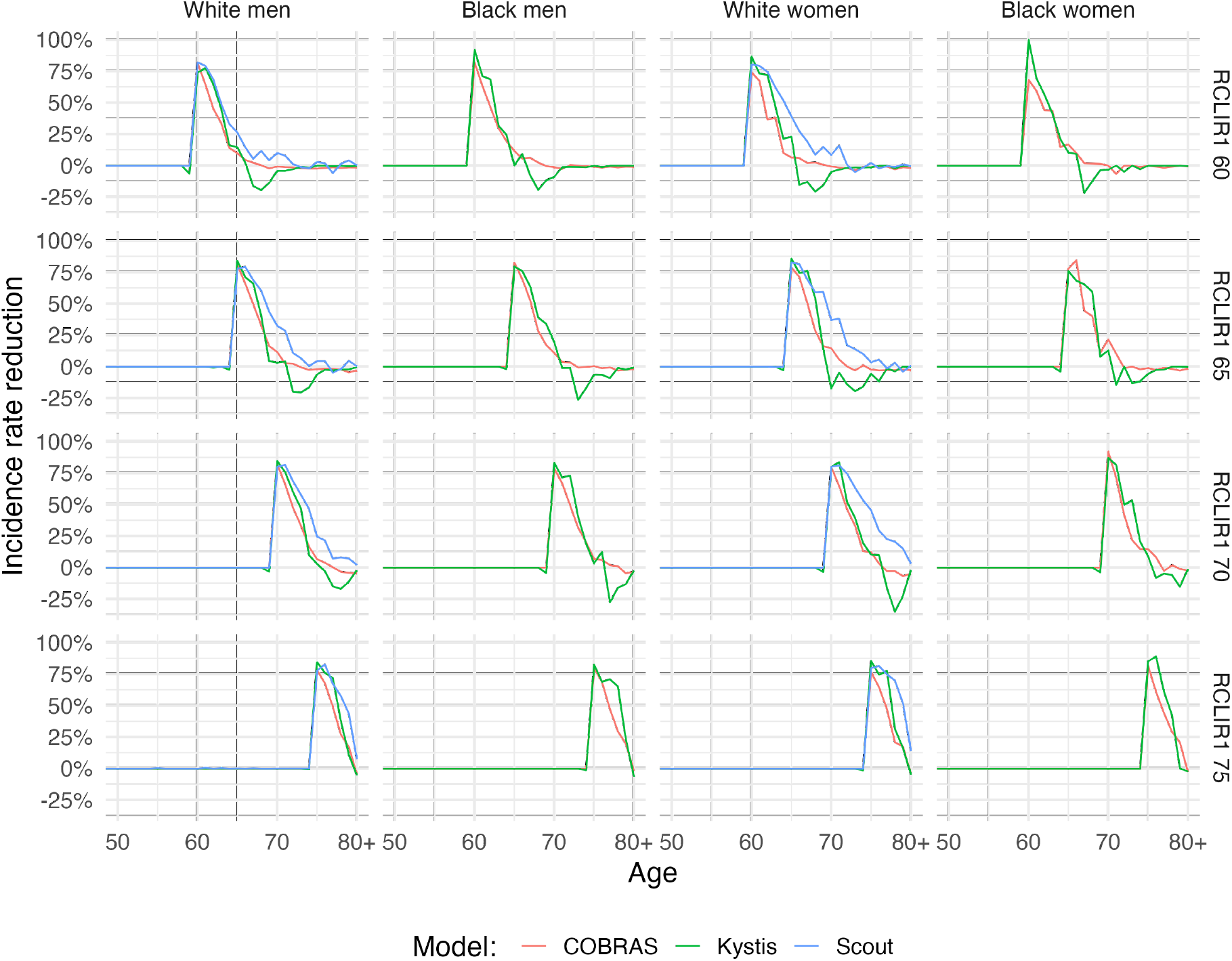
RCLIR-1 curves after one-time perfect screening by age, sex and race

### RCLIR-2 (realistic sensitivity, usual-care treatment)

Allowing usual-care treatment effectiveness further reduced clinical incidence gains (Figure 3). Compared with RCLIR-1, RCLIR-2 curves were lower and decayed more slowly, reflecting that some screen-detected cancers were not fully averted and could still progress to clinical detection despite earlier diagnosis. The incremental loss from RCLIR-1 to RCLIR-2 was largest in the model(s) with greater post-diagnosis recurrence/progression under usual care.

**Figure 3.**
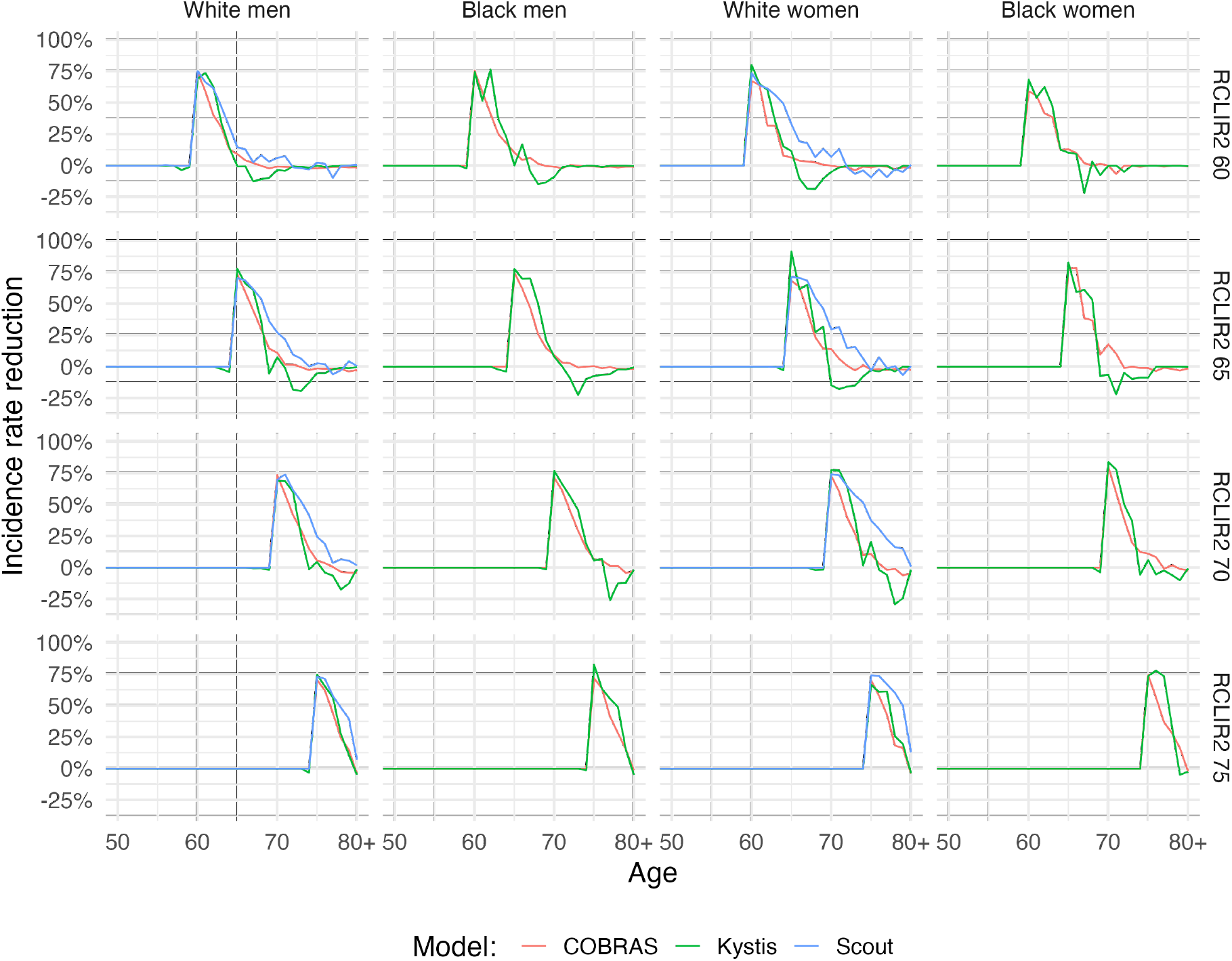
RCLIR 2 curves after one-time perfect screening by age, sex and race

Across models, the difference between MCLIR and RCLIR-1 quantifies missed preclinical disease due to imperfect detection at age 65, while the difference between RCLIR-1 and RCLIR-2 captures residual clinical incidence due to non-curative management. In all three models, the dominant reason for post-screening clinical incidence at older ages was new lesion emergence, not failures of detection or treatment (Figure 4). This pattern was strongest in COBRAS, consistent with its shorter preclinical sojourn time and faster replenishment of the detectable reservoir.

## Discussion

Using the CISNET MCLIR/RCLIR stress-test framework, we compared three independently developed bladder cancer microsimulation models under harmonized one-time screening counterfactuals. All models predicted that a single perfect screen would yield an immediate reduction in subsequent *clinically detected* incidence followed by attenuation toward zero as new lesions arise after the screening age, consistent with the theoretical behavior of MCLIR curves.

However, the size and persistence of the reductions differed somewhat across models. These differences align with emergent natural-history quantities: COBRAS produced shorter preclinical sojourn times from screen detectability to clinical diagnosis compared to Kystis and SCOUT. As a result, COBRAS implies a smaller detectable preclinical reservoir at age 65 and a faster post-screening replenishment of clinically detected disease, yielding smaller, less persistent MCLIR and RCLIR gains. Kystis and SCOUT imply a larger and longer-lived detectable reservoir, translating to larger and more sustained reductions.

Across all three models, the MCLIR curves declined relatively quickly after the screening age, indicating that, in the general population, the detectable preclinical phase of bladder cancer is limited in duration. Even under perfect detection and universally curative treatment, much of the clinical incidence at older ages arises from lesions that initiate after screening rather than from lesions missed or inadequately treated at screening.

This finding provides a mechanistic explanation for why population-wide, one-time screening is unlikely to produce large or durable reductions in clinical incidence. In practice, realistic screening (RCLIR) further lowers the achievable benefit, because imperfect sensitivity necessarily leaves part of the preclinical reservoir undetected at the screening age. Together, the paired MCLIR/RCLIR results suggest that any meaningful screening strategy for bladder cancer would likely need to be (i) repeated over time, (ii) targeted to groups with higher preclinical prevalence and/or longer sojourn times, or (iii) paired with tests that detect much earlier preclinical disease than cystoscopy alone.

The cross-model ordering and shapes we observed parallel insights from MCLIR applications in other CISNET cancer sites. The MCLIR method was designed explicitly to reveal whether models’ projected screening impacts are constrained chiefly by the detectable sojourn time and reservoir size implied by their natural-history assumptions ^9-12^. In cervix and lung comparisons, shorter implied preclinical sojourn times similarly yielded smaller and faster-decaying MCLIR curves, while longer sojourn times produced larger, more persistent reductions. ^11,12^. Our bladder cancer results therefore fit the broader CISNET pattern: when MCLIR is modest and transient, the natural history itself places a ceiling on screening effectiveness, regardless of downstream treatment assumptions.

From a policy standpoint, the results caution against expectations of large incidence reductions from single-round, general-population bladder cancer screening. Instead, they support exploration of *risk-stratified* screening or early-detection approaches, such as focusing on long-term heavy smokers, older men, or individuals with occupational exposures, where the detectable preclinical reservoir at a given age could be larger.

The results also underscore the value of tests that can detect earlier or smaller lesions (e.g., urine-based biomarkers) and/or strategies with *repeat* testing intervals tuned to the short sojourn times implied by the models. MCLIR/RCLIR provides a direct way to evaluate whether candidate tests meaningfully expand the detectable reservoir and thereby shift the achievable incidence-reduction ceiling.

The divergence in MCLIR magnitude across models points to specific empirical gaps that most strongly influence screening projections. The largest drivers are the timing of lesion emergence, and the interval from detectability to clinical diagnosis, quantities that are not directly observed and instead inferred through calibration. Comparative modeling thus helps prioritize data collection that would most shrink uncertainty in screening benefit, such as longitudinal studies of high-risk cohorts with repeated cystoscopy/biomarker testing to estimate detectable-phase duration; better characterization of symptom onset and diagnostic delay by stage/grade; and harmonized evidence on cystoscopy and emerging biomarker sensitivity for preclinical and very early NMIBC.

This study has several limitations. First, we evaluated a single birth cohort born in 1950; repeated rounds could yield different results. Second, the MCLIR scenario intentionally assumes perfect cure, and RCLIR-1 assumes universal cure after screen detection; these are not realistic clinical pathways but are necessary to decompose limits attributable to natural history versus management. Third, in RCLIR2 we assumed that the treatment effect is immediate. The standard of care for NMIBC is TURBT which removes most lesions at this stage. However, in practice, TURBT is generally done after the initial cystoscopy. Finally, all three models are calibrated to SEER 2010 incidence and stage distributions, so any registry misclassification or time-varying diagnostic practices could propagate into inferred sojourn times and reservoirs.

## Conclusion

In a standardized CISNET MCLIR/RCLIR comparison, COBRAS, Kystis, and SCOUT all imply that the detectable preclinical phase of bladder cancer is relatively short, placing a modest upper bound on the effectiveness of one-time screening in the general population. Cross-model differences in that bound are explained by differences in emergent sojourn times and progression speeds. These results provide a screening-relevant summary of bladder cancer natural history, clarify why screening projections may diverge, and motivate future work on risk-targeted and repeated early-detection strategies, supported by new empirical data to better resolve preclinical sojourn time and detectability.

## Data Availability

All data produced in the present study are available upon reasonable request to the authors

## Acknowledgements

The COBRAS and Kystis models were developed in the context of the Cancer Incidence and Surveillance Modeling Network (CISNET) program of the National Cancer Institute (NCI) of the National Institutes for Health (U01CA265750, principal investigators [PIs] Trikalinos and Jalal), and SCOUT is an NCI-funded affiliate research effort (R01CA262375, PI Kang). Together they comprise the CISNET Bladder Cancer Incubator Site. The results described here are based on COBRAS version 1.0.0, Kystis version 0.1.7.9004 and SCOUT version 1.0.0.

